# Screening for psychotic experiences and psychotic disorders in general psychiatric settings: a systematic review and meta-analysis

**DOI:** 10.1101/2024.04.14.24305796

**Authors:** Jacqueline A. Clauss, Cheryl Y. S. Foo, Catherine J. Leonard, Katherine N. Dokholyan, Corinne Cather, Daphne J. Holt

## Abstract

**Background:** The absence of systematic screening for psychosis within general psychiatric services contribute to substantial treatment delays and poor long-term outcomes. We conducted a meta-analysis to estimate rates of psychotic experiences, clinical high-risk for psychosis syndrome (CHR-P), and psychotic disorders identified by screening treatment-seeking individuals to inform implementation recommendations for routine psychosis screening in general psychiatric settings.

**Methods:** PubMed and Web of Science databases were searched to identify empirical studies that contained information on the point prevalence of psychotic experiences, CHR-P, or psychotic disorders identified by screening inpatient and outpatient samples aged 12-64 receiving general psychiatric care. Psychotic experiences were identified by meeting threshold scores on validated self-reported questionnaires, and psychotic disorders and CHR-P by gold-standard structured interview assessments. A meta-analysis of each outcome was conducted using the Restricted Maximum Likelihood Estimator method of estimating effect sizes in a random effects model.

**Results:** 41 independent samples (k=36 outpatient) involving n=25,751 patients (58% female, mean age: 24.1 years) were included. Among a general psychiatric population, prevalence of psychotic experiences was 44.3% (95% CI: 35.8-52.8%; 28 samples, n=21,957); CHR-P was 26.4% (95% CI: 20.0-32.7%; 28 samples, n=14,395); and psychotic disorders was 6.6% (95% CI: 3.3-9.8%; 32 samples, n=20,371).

**Conclusions:** High rates of psychotic spectrum illness in general psychiatric settings underscore need for secondary prevention with psychosis screening. These base rates can be used to plan training and resources required to conduct assessments for early detection, as well as build capacity in interventions for CHR-P and early psychosis in non-specialty mental health settings.

## INTRODUCTION

Psychotic disorders, such as schizophrenia, typically emerge during late adolescence or early adulthood, and are leading causes of disability.^1^ For those with psychotic disorders, a longer duration of untreated psychosis (i.e., the length of time that psychotic symptoms remain untreated)^2^ is associated with a poorer clinical course (greater symptom levels and lower treatment response rates) and worse long-term functional outcomes.^3–5^ Conversely, intervening early during the initial phase of psychotic illness (the first episode or shortly thereafter) has been shown to improve recovery outcomes.^6^ Thus, multiple lines of evidence indicate that it is important to identify and treat psychotic disorders as early as possible.

Consistent with this evidence supporting early intervention in psychosis, some studies have also shown that interventions in individuals who are at risk for developing a psychotic disorder due to the presence of subclinical symptoms of psychosis are valuable in reducing associated distress.^7^ Subclinical psychotic symptoms, often referred to as psychotic experiences (PEs), are relatively common in the general population (with ∼7% prevalence)^8^ and are frequently comorbid with non-psychotic psychiatric conditions, such as mood and anxiety disorders.^9^ The presence of PEs in those diagnosed with psychiatric conditions is associated with a number of adverse outcomes including more severe psychiatric symptoms,^10^ and increased risk of suicidal behaviors and mortality.^11,12^ Psychotic experiences are also associated with four-fold increased likelihood of developing a psychotic disorder^13^ and a three-fold increased risk of developing a psychiatric illness in general, including an increased risk for developing anxiety disorders, mood disorders, and substance use disorders.^14^ Thus, based on these data, earlier conceptualizations of transdiagnostic PEs as being beni^17^gn and representing “false positives” have been revised, given their association with poor clinical and functional outcomes.

On the more symptomatic end of the spectrum are those individuals who have PEs and evidence for symptomatic worsening over time, or a recent functional decline and genetic risk for a psychotic disorder, meeting the criteria for a clinical high-risk for psychosis syndrome (CHR-P).^15^ CHR-P is diagnosed using a standardized clinical interview.^16–18^ CHR-P was initially defined as a risk syndrome associated with elevated risk specifically for psychotic disorders. However, more recent evidence suggests that in addition to a 20-fold increased risk of developing a psychotic disorder,^19^ individuals with CHR-P have an increased risk for developing a broad range of psychiatric outcomes in the longer-term.^20^ Intervention studies of CHR-P have had mixed success,^21^ but generally support the value of treating this condition, given the distress that is typically associated with it. Thus, specialty clinics focusing on CHR-P patients have arisen in 15 countries to both address the current clinical needs and distress of these patients and to mitigate future risk for illness and functional impairment in this population.^22^ Thus, taken together, the convergent lines of evidence for the benefits of early detection and intervention in help-seeking individuals with clinical or subclinical psychotic symptoms highlight the need for effective screening approaches that can facilitate the prompt detection of psychotic disorders and PEs in general mental health settings. Such screening is a critical step towards prevention of poor long-term outcomes for individuals with these symptoms who are already seeking care for mental health concerns.

Despite the importance of early identification of psychotic symptoms and psychotic disorders, screening for psychotic symptoms is not routine in mental health settings and is even rarer in primary care. In contrast, suicide risk screening is required by Medicare^23,24^ and screening for substance use,^25^ depression^26^ and anxiety^26,27^ is recommended by the United States Preventive Services Task Force in primary care. An estimated 90% of those with a psychotic disorder diagnosis in the United States present to general outpatient mental health services two years prior to receiving a psychotic disorder diagnosis,^28^ and outpatient mental health providers are most frequently identified as the first clinical encounter for help-seeking CHR-P individuals.^29^ However, there are long referral and treatment delays for individuals with psychotic disorders ^30,31^. This may be related to the low confidence expressed by mental health professionals in their ability to identify psychotic disorders.^32^ However, implementing systematic screening for psychosis in outpatient mental health services is feasible and can reduce the duration of untreated psychosis.^33^

Knowledge of the prevalence of psychotic symptoms and disorders, along with the psychometric properties of potential screening measures (i.e., sensitivity, specificity), is necessary for estimating the number of people needed to screen to identify a true positive case and can also inform the design of screening and triaging protocols for identifying cases in real-world systems of mental healthcare. Secondary and tertiary prevention efforts can be built using such screening protocols, with the goal of preventing the onset of more severe illness or poor functional outcomes in individuals who are already suffering from psychiatric conditions. While prevalence rates of psychotic symptoms and psychotic spectrum disorders are known for the general population^34–36^ and in educational^37,38^ and primary care settings,^39^ little is known about the prevalence of psychotic disorders, PEs, and CHR-P in patients seeking treatment in general mental health settings. To our knowledge, there are no existing meta-analyses of rates of psychotic experiences and psychotic disorders in general psychiatric settings, although recent evidence suggests the CHR-P syndrome is common in patients presenting to psychiatric care.^40^

### Objectives

Therefore, we aimed to 1) provide an estimate of the prevalence rates of psychotic experiences, CHR-P, and psychotic disorders in general psychiatric settings; 2) test for the effects of demographic variables and publication-related variables on prevalence rates, 3) summarize the psychometric properties of recommended screening measures; and 4) make recommendations for implementing psychosis screening in general mental health settings.

## METHODS

We conducted a systematic review and meta-analysis following the PRISMA guidelines^41^ to calculate the overall prevalence of psychotic experiences, CHR-P syndrome, and psychotic disorders based on screenings conducted in general psychiatry settings. General psychiatric settings were defined as community, outpatient, inpatient, and/or emergency settings providing non-specialist psychiatric, psychological, or psychosocial care. Primary medical settings and specialized psychosis programs or services were excluded.

### Search strategy

Medical and social science electronic databases (Web of Science and PubMed) were searched to include studies published from January 1, 1990, to July 1, 2023. Non-peer reviewed manuscripts, theses, conference proceedings, protocols, and abstracts were not included. The references of relevant studies were reviewed to identify additional studies. Searches were based on a combination of controlled vocabulary index terms related to: 1) psychotic experiences or psychotic disorders (e.g., psychosis OR psychotic OR schizophrenia); 2) screening and detection (e.g., screen* OR detect* or identif*); and 3) general psychiatric settings (e.g., help-seeking population OR psychiatric population OR outpatient mental health OR community mental health OR inpatient mental health). Filters were used to exclude studies based on *a priori* exclusion criteria (e.g., studies on screening in older adults (ages 65+), school/primary care/traditional healer settings). See Supplemental File 1 for the full list of search terms used.

### Eligibility criteria

Peer-reviewed, empirical studies written in English were eligible for inclusion if they contained original, quantitative data on point prevalence rates of psychotic experiences, psychotic disorders or CHR-P identified by screening treatment-seeking samples aged 12-64 receiving general psychiatric care. Studies were included if they used either: 1) an established/validated structured clinical interview to assess the presence of psychotic disorders diagnoses (schizophreniform disorder, schizophrenia, schizoaffective disorder, presence of psychotic symptoms in psychosis risk syndrome interview) or a psychosis-risk syndrome (attenuated psychotic symptoms syndrome, brief intermittent psychotic symptoms syndrome, or genetic risk and deterioration syndrome), or 2) a validated questionnaire to identify psychotic experiences. Studies were not excluded based on country of origin or sample size.

We excluded publications describing psychosis screening in samples that included: a) older adults, due to the diagnostic complexity and comorbidity of psychotic symptoms with other medical and neurodegenerative conditions (e.g., dementia); b) school-based settings or populations (e.g., school counseling centers), medical primary care settings, specialty psychosis detection and evaluation centers (e.g., CHR-P clinics), medical inpatient or medical emergency room settings, and traditional/indigenous healer settings; and c) criminal legal system-involved, forensic, veteran populations due to their unique healthcare systems. We excluded publications which reported on screening for biomarkers or genetic markers of psychosis risk. Unpublished theses, protocols, editorials, opinion pieces, qualitative studies, and studies without primary data (e.g., reviews) were excluded. Studies that did not use established psychometric instruments to detect psychotic symptoms and likely psychotic disorders (e.g., medical records review, insurance claims, non-structured clinical interview) were excluded. Overlapping studies, determined by the setting’s name (if available), country, participating authors, recruitment period, and instruments, were excluded. In the case of overlap, the report of the largest representative sample was selected.

### Screening and data extraction procedures

All identified references were managed in Covidence, a web-based collaboration software platform that streamlines the production of systematic and other literature reviews.^42^ All titles and abstracts were independently screened by at least two trained reviewers and the first authors (JAC and CYSF) resolved disagreements between coders and mutually agreed on studies eligible for full-text review. A double-screening approach was performed for subsequent eligibility screening and data extraction to avoid systematic and random errors.^43^ Full-texts were independently reviewed by JAC and CYSF, and JAC and CYSF resolved differences in final sample. Two trained reviewers (KD, CJL) independently extracted data from included studies using structured tables, including details on: author and year; country; clinical setting (inpatient, outpatient, child/youth mental health); sample size screened; demographic characteristics (age, % female, race/ethnicity, when available); outcomes assessed (psychosis symptoms, CHR-P, and/or psychotic disorders); screening measures used (including if there was a pre-screening tool); and screening prevalence rates of psychotic symptoms, psychotic disorders, and/or CHR-P syndromes. JAC and CYSF audited extracted data for accuracy; inconsistencies were minor and resolved through consensus discussion. Quality of study was assessed with the JBI Critical Appraisal Instrument for Studies Reporting Prevalence Data that included 8 criteria: appropriate sampling strategy, sample size, sample representativeness, response rate and attrition, validity of measures, and reliability and validity of screening procedure.^44^ We applied an *a priori* benchmark of N≥100 for adequate sample size. Included studies were rated as “potentially low-quality” if they did not adequately meet a minimum quality rating of 60% (i.e., 6/8).

Independent samples from included studies were categorized into three outcomes for meta-analysis, specifically, screening prevalence rates for 1) psychotic experiences, 2) CHR-P, or 3) psychotic disorders. Screening prevalence rates for psychotic experiences were based on the cut-off score the screening measure in the study used to determine positive screen for likely psychotic disorders. In samples with screening rates derived from multiple measures, reviewers (CYSF and JAC) came to a consensus on which measure was the most well-validated and selected the prevalence rate determined by that measure.

### Statistical analysis

For each sample and outcome, screening prevalence rates and their variances were calculated. A meta-analysis of combined screening prevalence rate for each outcome was estimated using the Restricted Maximum Likelihood (REML) Estimator method. The REML method uses random effects to estimate overall point prevalence and confidence intervals without requiring normal distributions and is robust to small sample sizes.^45,46^ The random effects model weights larger sample sizes with lower variance higher than smaller samples with higher variance within the meta-analysis. Meta-analyses were repeated with the following sensitivity analyses 1) excluding inpatient psychiatric samples that could potentially inflate prevalence rates of psychotic disorders, and 2) excluding studies with potentially low-quality data. Additional analyses assessing child and adolescent clinic samples were also conducted. Meta-regression analyses to determine the effects of publication year, sex (% female in sample), and age (sample mean/median, or median of study’s reported age range) on overall meta-analytic prevalence rates for each outcome were tested. Two-sided statistical tests and a significance level of alpha = .05 were used. Meta- and moderator analyses were performed using the Metafor package in R Studio (Version 2023.09.1+494).^46^ Egger’s test for publication bias was performed. For analyses with significant effects of publication bias, analyses were repeated using a trim and fill method to impute missing datapoints.

## RESULTS

The search strategy yielded a total of 5,525 publications on Web of Science and 553 on PubMed. Citation searching revealed an additional 20 publications for potential inclusion. Forty studies published from 2006 to 2023 with 41 independent samples (total N = 25,751; 58% female, mean age: 24.1 years) met eligibility criteria for the systematic review (see Figure 1 for CONSORT flow diagram). Five studies had potentially low-quality data.

**Figure 1.**
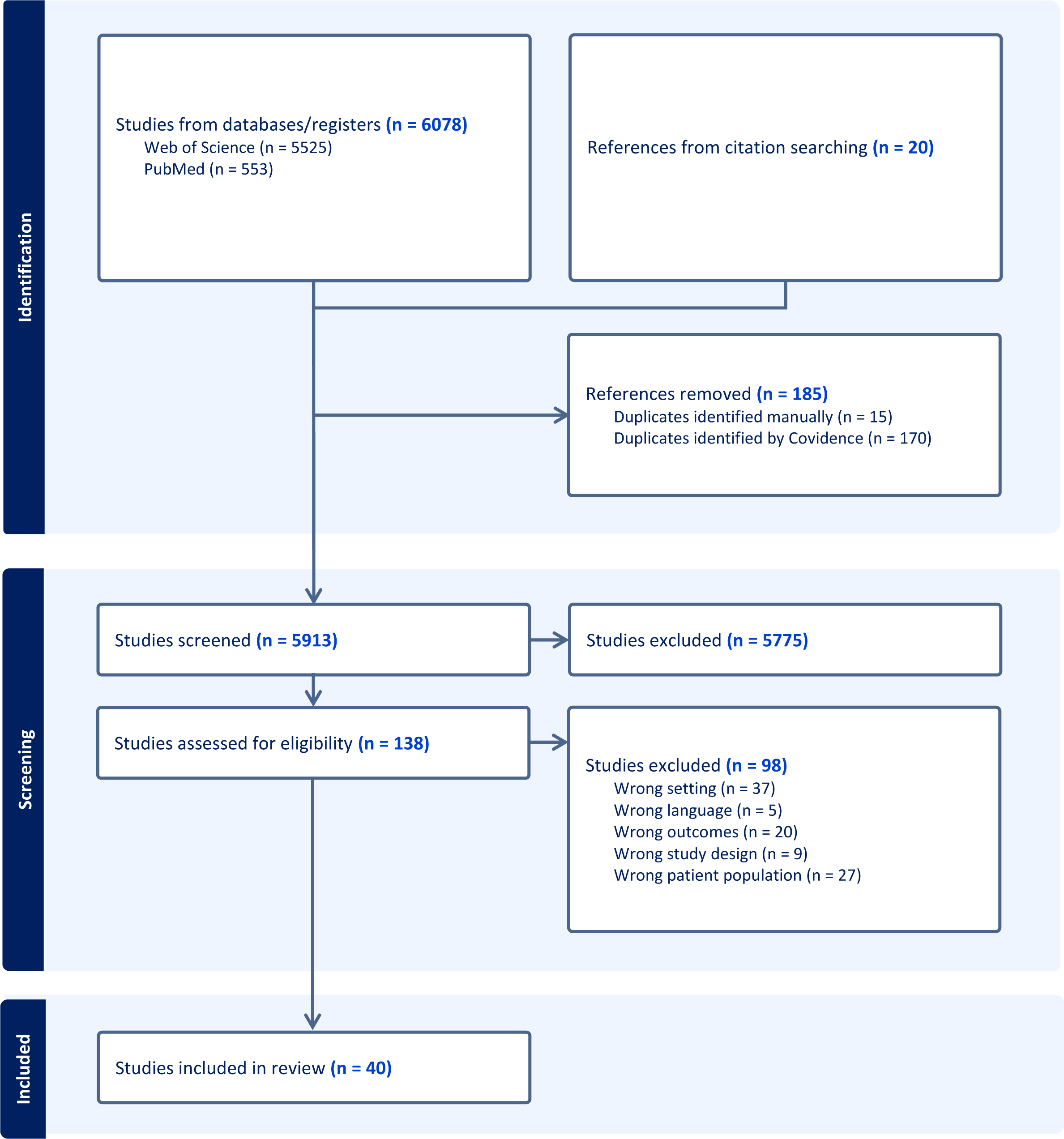
CONSORT flow diagram of systematic literature review.

Table 1 summarizes the 41 independent outpatient (k=36), inpatient (k=2), and combined inpatient and outpatient (k=3) screened samples across 13 different countries. Two samples were recruited from child and adolescent inpatient units, and 14 from child, adolescent, or young adult outpatient mental health settings serving patients with variable age ranges, from 6-to 24-years-old. The majority of included studies aimed to either determine prevalence of psychotic experiences or psychosis-risk syndromes (k=6), or to examine screening properties of self-report instruments and their concordant validity against other diagnostic assessments or clinical measures (k=8). Several studies consisted of evaluations of screening methods implemented as part of research studies or quality improvement initiatives (k=9). Twenty-eight (28) samples were screened for prevalence of psychotic experiences (16 adult outpatient samples, 8 child and adolescent outpatient samples, 2 adult combined inpatient and outpatient samples, 1 inpatient adult sample, and 1 combined child and adolescent inpatient and outpatient sample), 28 samples were screened for CHR-P (14 adult outpatient samples, 9 child and adolescent outpatient samples, 2 adult combined inpatient and outpatient samples, 1 inpatient adult sample, and 1 combined child and adolescent inpatient and outpatient sample), and 32 samples were screened for psychotic disorders (18 adult outpatient samples, 10 child and adolescent outpatient samples, 2 adult combined inpatient and outpatient samples, 1 inpatient adult sample, and 1 combined child and adolescent inpatient and outpatient sample).

**Table 1.**
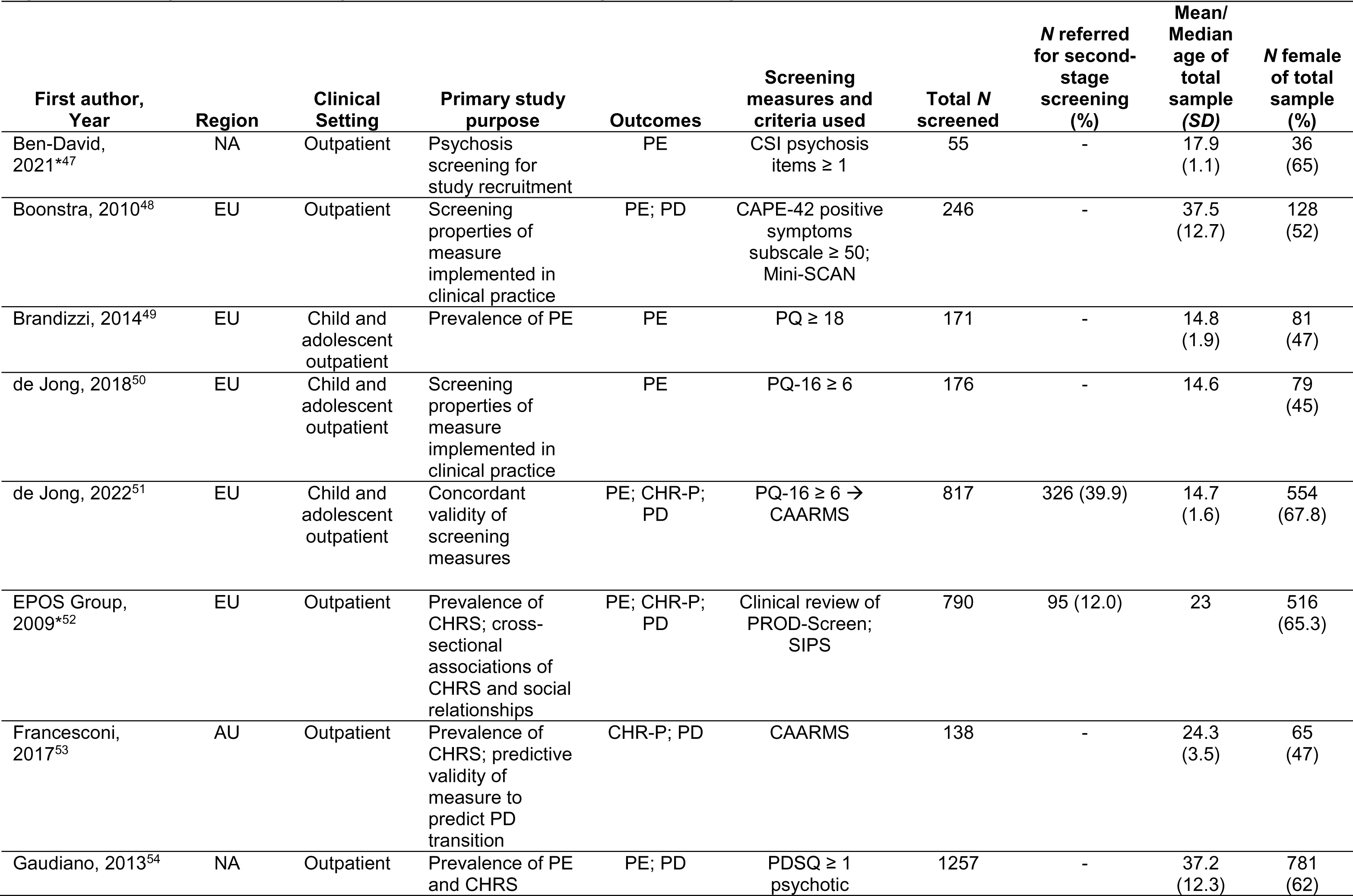
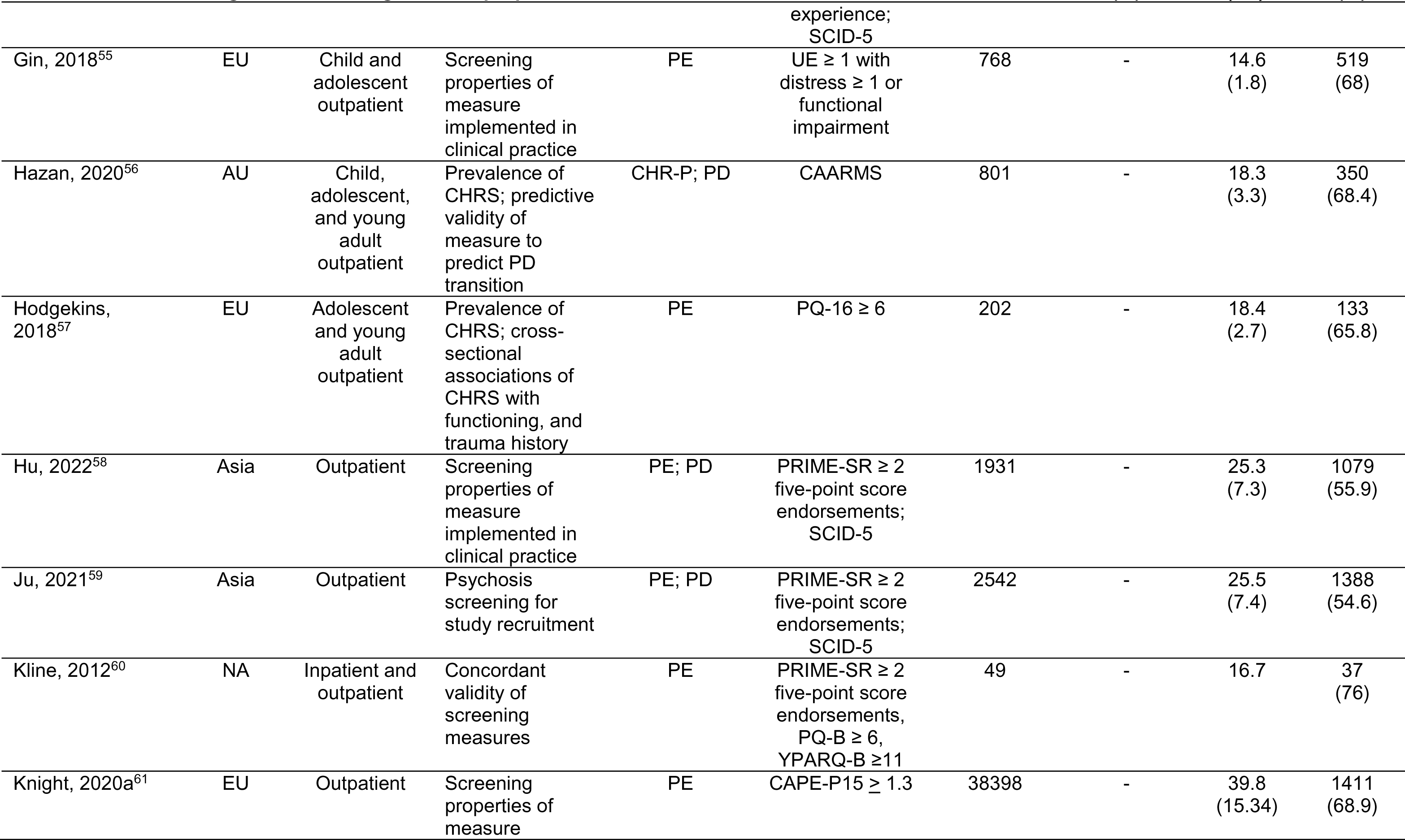
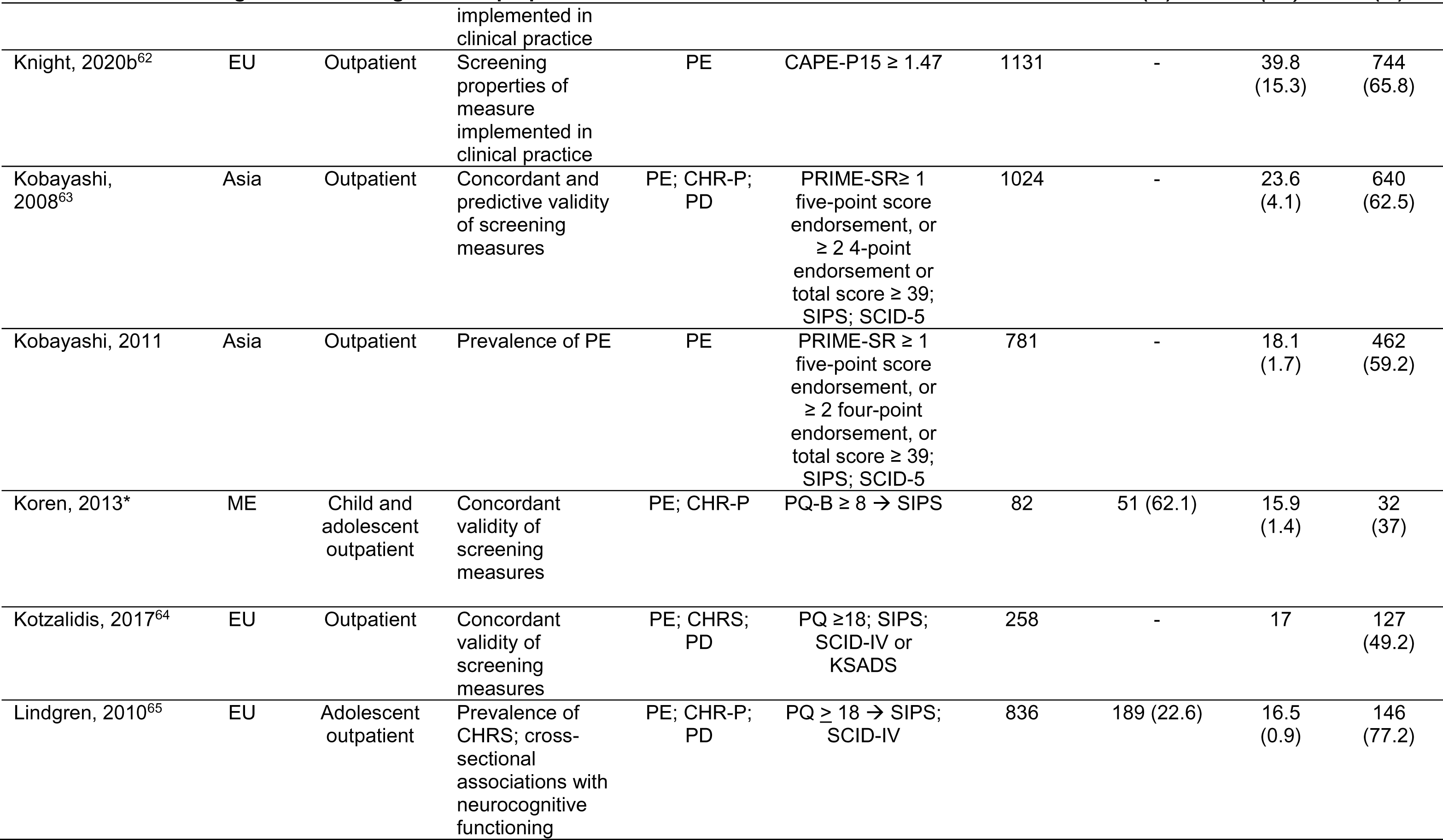
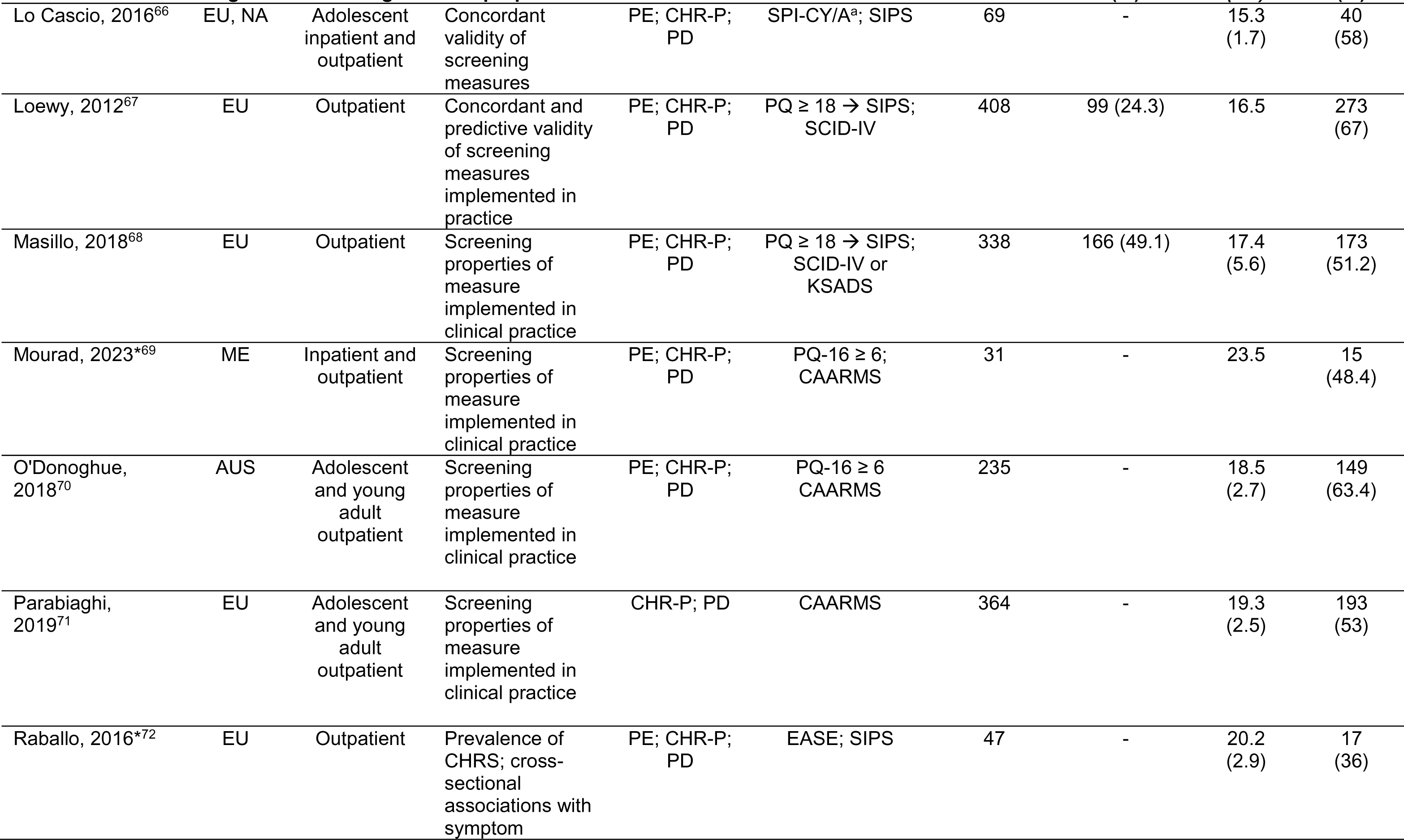
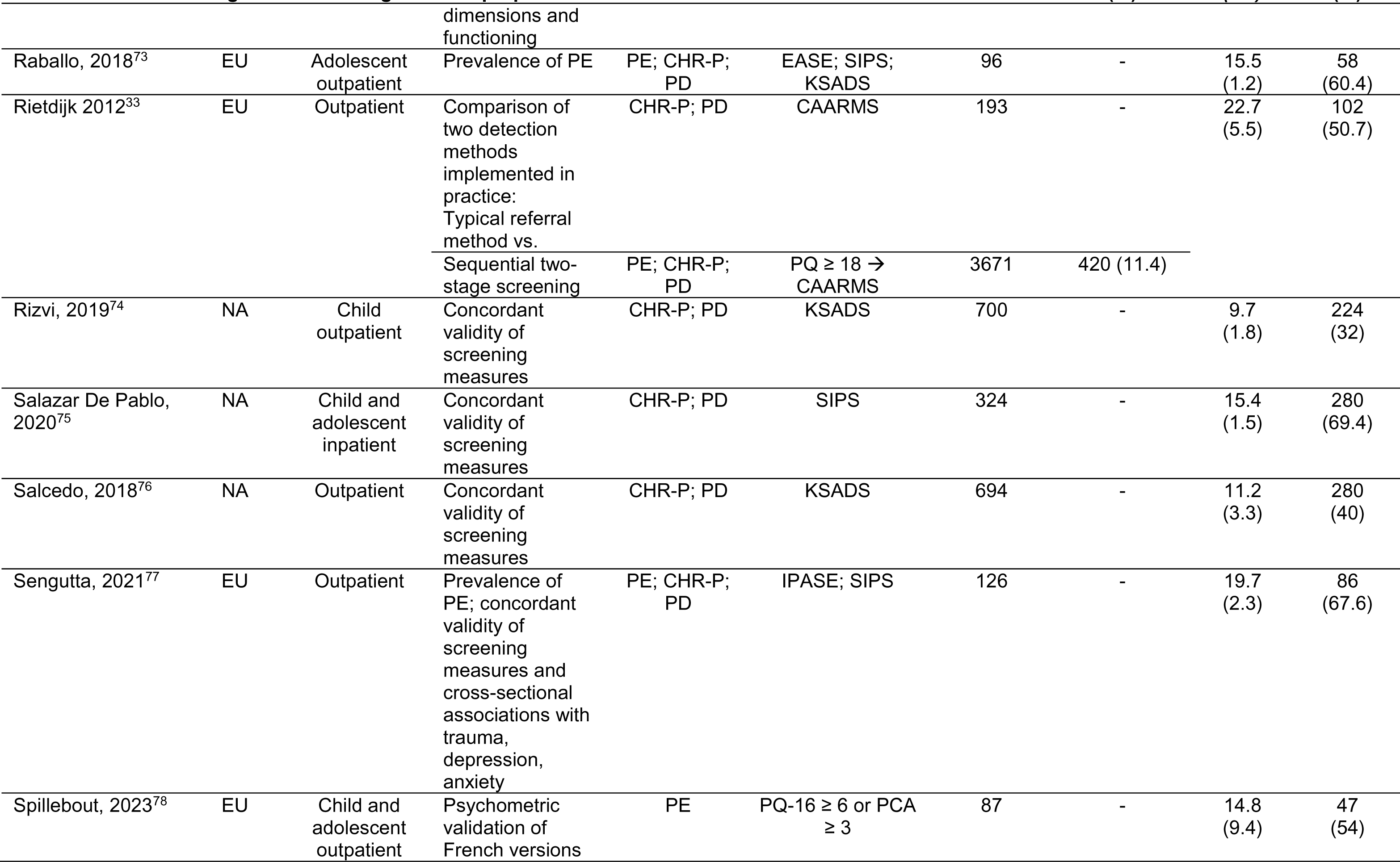
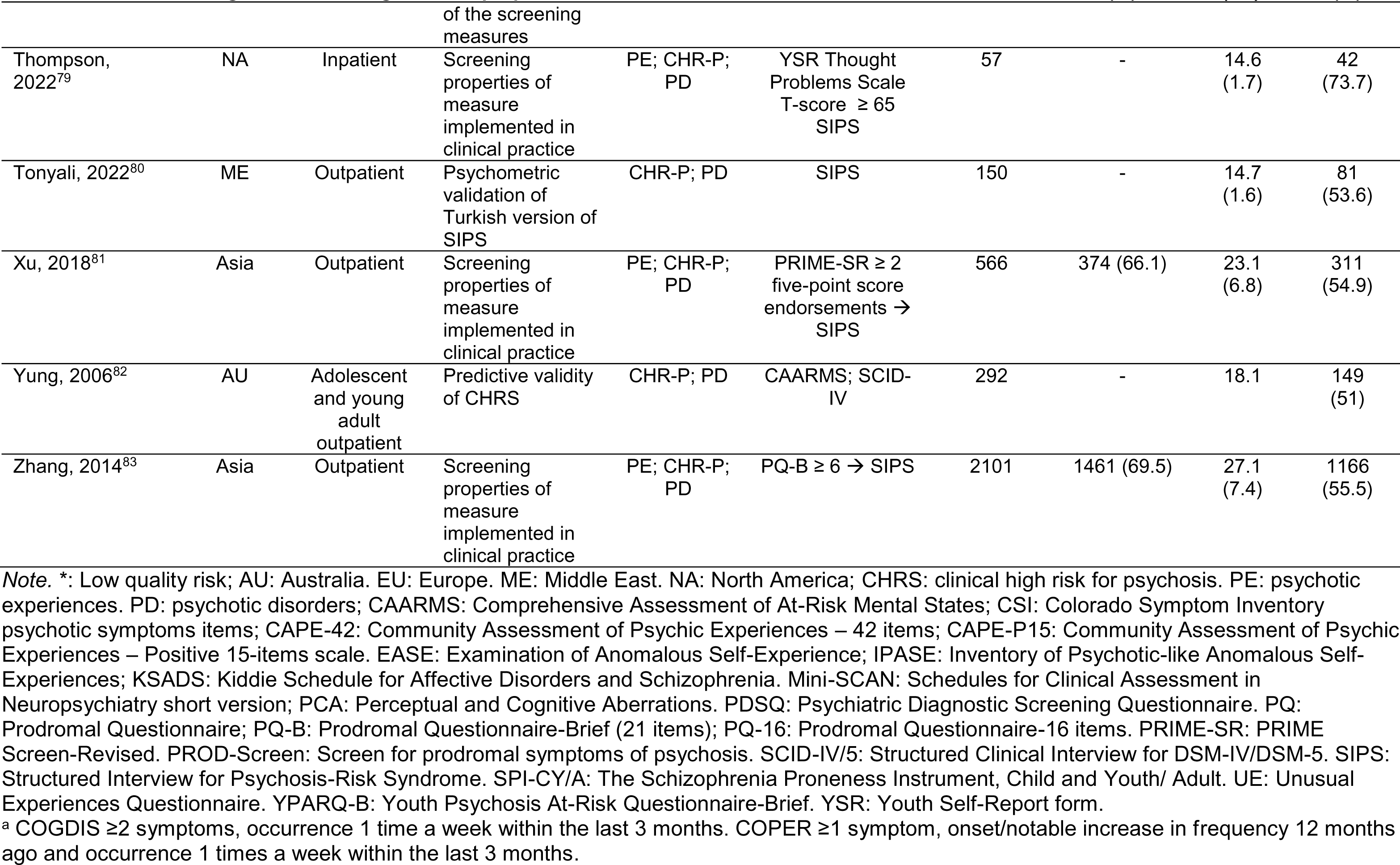
Summary of studies in general psychiatric settings identifying clinically indicated psychotic symptoms, clinical high risk for psychosis, and psychotic disorders through screening.

### Screening measures

A total of 15 established self-report questionnaires and one parent-/teacher-report questionnaire, some of which were translated and validated in the local language, were used to assess psychotic experiences. Table 2 summarizes the screening properties of the more frequently used measures for each outcome among the included studies, as well as additional measures with strong validity that are brief and recommended for general psychiatric settings (see Discussion).

**Table 2.** Recommended screening and assessment measures for psychotic spectrum symptoms and disorders in general psychiatric settings.

| Psychotic Symptoms |  |  |  |  |  |  |  |
| --- | --- | --- | --- | --- | --- | --- | --- |
| Measure | Format and access <sup>a</sup> | Number of items (estimated assessment time in minutes) | Age range in validation study (years) | Psychotic symptom domains assessed | Suggested threshold for clinically indicated psychotic symptoms in general psychiatric settings | Sensitivity predicting CHR-P status using suggested threshold <sup>b</sup> | Specificity predicting CHR-P status using suggested threshold <sup>b</sup> |
| Prodromal Questionnaire (PQ)<sup>84</sup> | Self-report | 92 (20) | 12-35 | Positive; Negative; Disorganized; General psychiatric; Distress from symptoms | ≥18 positive symptoms | 80-91%<sup>85</sup> | 40-90%<sup>85</sup> |
| Prodromal Questionnaire-Brief (PQ-B)<sup>86</sup> | Self-report | 21 (10) | 12-17 | Positive; Distress or impairment from symptoms | ≥6-7 total symptoms or ≥24 total distress score | 82-84%<sup>85</sup> | 60-64%<sup>85</sup> |
| Prodromal Questionnaire-Brief Child Version (PQ-BC)<sup>87</sup> | Self-report | 21 (10) | 9-10 | Positive; Distress from symptoms using a visual scale | NA | NA | NA |
| Prodromal Questionnaire-16 (PQ-16)<sup>88</sup> | Self-report | 16 (5) | 18-35 | Positive; Negative | ≥6 total symptoms or ≥8 total distress score | 78-90%<sup>85</sup> | 54-91%<sup>85</sup> |
| Prevention through Risk Identification, Management, and Education (PRIME) Screen-Revised (PS-R)<sup>89</sup> | Self-report | 12 (5) | 16-30 | Positive; Loss of insight | ≥1 item scored at 6 or ≥2 items scored at 5 or ≥39 total score | 100% | 74% |
| Youth Psychosis at Risk Questionnaire-Brief (YPARQ-B)<sup>90</sup> | Self-report | 28 (10) | 14-18 | Positive | ≥11-13 total symptoms | 65-100%<sup>60</sup> | 76-80%<sup>60</sup> |
| PROD-Screen<sup>91</sup> | Self-report or interview screen | 21 (10) | 14+ | Positive; Negative; Disorganized; and General psychiatric symptoms | ≥2 total symptoms | 80% | 75% |

| Adolescent Psychosis Symptom Screener (APSS)<sup>92</sup> | Self-report | 7 (5) | 11-13 | Positive | ≥1 symptom, with endorsing auditory hallucinations having the highest predictive power | 20-70% | 87-100% |
| --- | --- | --- | --- | --- | --- | --- | --- |
| 2-item psychosis screener<sup>93</sup> | Self-report | 2 (1) | 12-25 | Positive | ≥1-2 symptoms | 32-59% | 68-95% |
| **Clinical High Risk for Psychosis** |  |  |  |  |  |  |  |
| Measure | Format and access | Estimated assessment time (mins) | Age range in validation study (years) | Measures and symptom domains assessed | CHR-P syndromes and diagnoses assessed | Sensitivity of CHR-P criteria predicting onset of psychotic episode <sup>c</sup> | Specificity of CHR-P criteria predicting onset of psychotic episode <sup>c</sup> |
| Structured Interview for Psychosis-Risk Syndromes (SIPS)<sup>94</sup> | Semi-structured interview by certified assessors; 1-2 day-long training | 60-120 | 12-45 | Family history of mental illness; Positive, Negative, and Disorganized prodromal psychotic symptoms and general psychiatric symptoms; Global Assessment of Functioning scale; Schizotypal personality disorder criteria checklist | Presence of psychotic syndrome; brief intermittent psychotic syndrome (BIPS); attenuated positive symptom syndrome (APSS); genetic risk and functional deterioration syndrome (GRD) | 100% | 71% |
| Comprehensive Assessment of At-Risk Mental States (CAARMS)<sup>95</sup> | Semi-structured Interview by mental health professionals | 60-120 | 15-29 | Disorders of thought content; perceptual abnormalities; conceptual disorganization; motor changes; concentration and attention; emotion and affect; subjectively impaired energy and impaired tolerance to normal stress; social and occupational<br>functioning<br>assessment scale | Attenuated Positive Symptom Intensity and Frequency (APS); Brief Limited Intermittent Psychosis Syndrome (BLIPS); Vulnerability group | 83% | 74% |
| Positive Symptoms and Diagnostic Criteria for the CAARMS Harmonized with the SIPS (PSYCHS)<sup>96</sup> | Semi-structured interview by trained assessors | 60-120 | 12-30 | Attenuated delusions; attenuated hallucinations; attenuated thought disorder | Harmonized criteria for CAARMS/SIPS psychosis diagnosis and CHR-P syndrome criteria | NA | NA |
| Mini-SIPS<sup>97</sup> | Free 30-minute online training; publicly available | 15-20 | 12-45 | Positive psychotic symptoms | DSM-5 Attenuated Psychosis Syndrome (APS) clinical diagnosis; SIPS attenuated positive symptom syndrome (APSS) progression | NA | NA |

| Psychotic Disorders |  |  |  |  |  |  |
| --- | --- | --- | --- | --- | --- | --- |
| Measure | Format and access | Estimated assessment time (mins) | Age range in validation study (years) | Measures and domains assessed | Diagnoses assessed |  |
| Schedule for Affective Disorders and Schizophrenia for School-Age Children present and lifetime version (KSADS-PL DSM-5)<sup>98</sup> | Semi-structured parent and child interview by mental health professional or clinically trained assessor; publicly available and free to use | 90-120 | 6-18 | Developmental history; current and past psychiatric symptoms; Children's Global Assessment Scale | DSM-5 Axis I child and adolescent psychiatric disorders including schizophrenia and schizoaffective disorder; Scores of 2 indicate sub-threshold presentation and scores of 3 indicate threshold presentation of symptoms |  |
| Computerized versions of the Kiddie Schedule for Affective Disorders and Schizophrenia (KSADS-COMP)<sup>99</sup> | Web-based child and parent self-administered, and clinician-administered | 50-60 | 6-18 | Developmental history; current and past psychiatric symptoms; Children's Global Assessment Scale | DSM-5 Axis I child and adolescent psychiatric disorders including schizophrenia and schizoaffective disorder; Scores of 2 indicate sub-threshold presentation and scores of 3 indicate threshold presentation of symptoms |  |
| Structured Clinical Interview for DSM-5 Clinician Version (SCID-5-CV)<sup>100</sup> | Semi-structured Interview by mental health professional or clinically trained assessor; publicly available and free to use | 45-120 | 18+ | Current psychiatric symptoms; Functioning | DSM-5 diagnoses |  |
| NetSCID<sup>101</sup> | Web-based version of the SCID-5; \$25 per clinician assessment | 30-45 | 18+ | Current psychiatric symptoms; Functioning | DSM-5 diagnoses | |

*Note.* CHR-P: clinical high risk for psychosis; NA: not currently available or reported in published studies.
<sup>a</sup> All screening measures for psychotic symptoms reported here are available in the public domain and free to use.
<sup>b</sup> Range of sensitivity and specificity of measure based in studies that screened treatment-seeking samples in general psychiatric settings using screening thresholds for clinically indicated psychotic symptoms. Measure's positive screen criteria were validated against gold-standard, clinical interview assessments of CHR-P (e.g., SIPS, CAARMS). Screening properties of measure based on studies, where measure was validated against gold-standard, clinical interview assessment of CHR-P.
<sup>c</sup> We reported predictive validity of the CHR-P assessment at 6 months from the original validation study. Specificity of CHR-P criteria predicting onset of psychotic episode increases with time.

The 92-item Prodromal Questionnaire (PQ) and its shorter versions (16-item PQ-16, 21-item PQ-Brief) were the most frequently used self-report screening measures for psychotic experiences (13/28 samples; 46%), followed by the PRIME Screen-Revised (PRIME-SR) (7/28; 25%). In general psychiatric settings, the concurrent validity of assessing psychotic experiences using the recommended cutoffs for PQ, PQ-16, PQ-B, or PRIME-SR against gold-standard CHR-P assessments was strong (sensitivity: 0.78 to 1.0; specificity: 0.54 to 0.91), indicating that these self-report measures can accurately distinguish between those likely to meet criteria for CHR-P syndrome and those who do not (Table 2). CHR-P status was most frequently determined using the Structured Interview for Psychosis-Risk Syndrome (SIPS; 16/28 samples with CHR-P screening; 57%), followed by the Comprehensive Assessment of At-Risk Mental States (CAARMS; 9/28, 32%), and the Kiddie Schedule for Affective Disorders and Schizophrenia (KSADS, 2/28, 7%). Psychotic disorder diagnosis was ascertained by structured clinical diagnostic interviews such as the KSADS or the Structured Clinical Interview for DSM-5 or DSM-IV (SCID-5/IV; 11/33), or meeting threshold for psychotic disorder through the SIPS or CAARMS assessments. Supplemental File 2 compiles a comprehensive list of known validated instruments for assessing psychosis symptoms and disorders, screening properties in general psychiatric settings, and languages in which they are available.

### Two-stage screening

Nine samples were screened for CHR-P using a sequential, two-stage, screening method, where an enriched sample meeting a threshold for the presence of psychotic experiences on a brief screening tool was followed up with gold-standard assessments of CHR-P. Seven (77.7%) samples utilized the PQ, PQ-16, or PQ-B as the initial screening measure and six followed up with the SIPS to determine CHR-P status. The EDIE-NL study^33^ was the only published study to our knowledge that compared the effectiveness of a two-stage screening strategy (PQ ≥ 18 followed by the CAARMS) to a traditional clinician referral followed by CAARMS in an outpatient help-seeking population. In this study, two-stage screening identified twice as many first-episode psychosis patients and three times as many CHR-P patients at entry to mental health services than clinician referral.

### Meta-analyses of point prevalence

See Figure 2 for forest plots of meta-analysis of rates identified by screening for each outcome. Heterogeneity was high for all outcomes (psychotic experiences: I^2^=99.6%; CHR-P: I^2^=99.3%). I^2^ represents the percentage of variability in the effect sizes which is not caused by sampling error (> 75% = high variability). I^2^ cannot be calculated for any studies for which variance is 0, thus could not be calculated for psychotic disorders. τ^2^ is a measure of between-study variance and provides an estimate of underlying distribution of true effect sizes (τ^2^ ± standard error (SE): psychotic experiences: 0.052 ± 0.014; CHR-P: 0.028 ± 0.008; psychotic disorders 0.008 ± 0.002). Heterogeneity remained high for sensitivity analyses of high-quality outpatient studies (psychotic experiences: I^2^=99.7%, τ^2^ ± SE: 0.053 ± 0.017; CHR-P: I^2^=99.5%, τ^2^ ± SE: 0.026 ± 0.009; psychotic disorders: I^2^=99.5%, τ^2^ ± SE: 0.009 ± 0.003).

**Figure 2.**
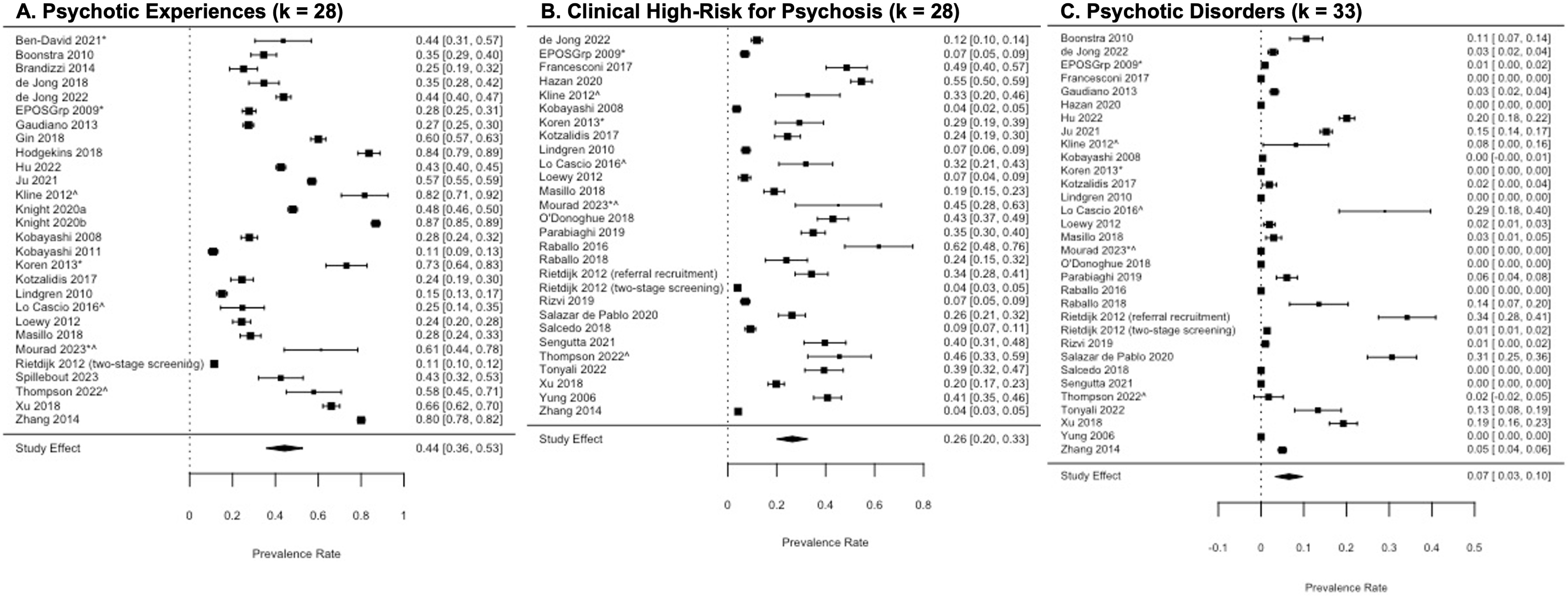
Meta-analyses of point prevalence of psychotic experiences, clinical high-risk for psychosis syndrome, and psychotic disorders in general psychiatric settings. **A. Overall prevalence rate of positive screen for psychotic symptoms in general psychiatric population.** Twenty-eight samples were identified including 21,957 participants and 9,294 met established screening criteria for threshold psychotic symptoms. Overall prevalence of threshold psychotic experiences detected was 44.3% [95% CI: 35.8-52.8%]. **B. Overall prevalence rate of clinical high-risk for psychosis syndrome identified by structured clinical interview in general psychiatric settings.** Twenty-eight (28) samples were identified including 14,395 participants who were systematically assessed for CHR-P and 1,937 met criteria for CHR-P. The overall prevalence rate for CHR-P detected by screening in the general psychiatric population was 26.4% [20.0-32.7%]. **C. Overall prevalence rate of psychotic disorders identified by structured clinical interview in general psychiatric settings.** Thirty-two (32) samples were identified including 20,371 participants who were systematically assessed for psychotic disorders and 1,391 met criteria for a psychotic disorder by standardized clinical interview. The overall prevalence rate for psychotic disorder detected by screening in the general psychiatric population was 6.6% [3.3-9.8%] in the general psychiatric population. *Indicates low quality data. ^Indicates inpatient or combined inpatient/outpatient sample.

#### Psychotic experiences

In 28 independent samples, 21,957 participants (55.8% female; mean age: 25.7 years) were screened for psychotic symptoms, and 9,294 met established screening criteria for psychotic experiences. In the general psychiatric setting, the prevalence of psychotic experiences detected by screening across all samples was 44.3% [95% CI: 35.8-52.8%].

Among the nine samples recruited from child and adolescent clinical programs, the prevalence rate of psychotic symptoms was 44.8% [29.5-60.0%]. In sensitivity analyses excluding four samples with inpatients (k=24), the overall prevalence rate was 42.4% [33.3-51.5%]. Similarly, high quality studies with outpatients (k=21) yielded an overall prevalence rate of 41.7% [31.8-51.6%].

There was a significant effect of publication year on psychotic experiences, with higher rates for psychotic experiences reported in more recent papers (β = 0.019, p = 0.05). There were no significant effects of age (β = 0.005, p = 0.31) or sex (β = 0.18, p = 0.67) on prevalence rates of psychotic experiences. In comparing the two most common scales used to assess psychotic experiences, the PRIME (k = 6) and the Prodromal Questionnaire (k = 14), inclusive of all versions of each, there was no significant effect of scale on rates of psychotic experiences (β = - 0.06, p = 0.66). There was no evidence of publication bias in samples reporting psychotic experiences (see Supplemental File 3, Egger’s test for publication bias: Z = 1.08, p = 0.28).

#### Clinical high-risk for psychosis

In 28 independent samples, 14,395 participants were systematically assessed for CHR-P (42.2% female; mean age: 14.1 years) and 1,937 met criteria for CHR-P. The overall prevalence rate for CHR-P detected by clinical interview in the general psychiatric population was 26.4% [20.0-32.7%]. Among 11 samples recruited from child and adolescent programs, the overall prevalence rate of CHR-P was 28.1% [18.9-37.3%]. Among the nine samples that utilized secondary screening, the overall prevalence rate was lower, 11.5% [11.4-16.5%]. In sensitivity analyses excluding inpatient samples (k=24), the overall prevalence rate was 24.6% [17.7-31.5%]; consistent with this, high quality studies conducted in outpatient samples (k=21) had an overall prevalence rate of 23.7% [16.7-30.7%].

Higher rates of CHR-P were reported in more recently published papers; however, this effect was marginally significant (β = 0.015, p = 0.06). There were no significant effects for age (β = 0.008, p = 0.38) or sex (β= -0.08, p = 0.78) on CHR-P rates. In comparing the two most used interviews for CHR-P, the CAARMS (k = 9) and the SIPS (n = 15), there was no significant effect of interview type on CHR-P rates (β = -0.098, p = 0.16). In overall analysis, there was significant evidence of publication bias, such that smaller samples provided higher estimates of point prevalence (see Supplemental File 3, Egger’s test for publication bias: β = 0.082 [0.01-0.15], Z = 5.93, p < .0001); however, trim and fill analysis did not change the results, as no left-sided studies were imputed.

#### Psychotic disorders

In 32 independent samples, 20,371 participants were assessed for psychotic disorders using structured clinical diagnostic interviews only, (55.6% female; mean age: 22.6 years) of whom 1,391 patients met criteria for a psychotic disorder. Psychotic disorders had a prevalence of 6.6% [3.3-9.8%] in the general psychiatric population. High quality studies with outpatients (k=25) had an overall prevalence rate of 7.1% [3.4-10.8%]. Among the 11 samples recruited from child and adolescent programs, the overall prevalence rate of psychotic disorders was 6.9% [4.4-13.4%]. For the nine samples which utilized sequential screening strategies, the overall prevalence rate was lower, 3.7% [0.0-7.4%].

There was a significant effect of publication year (β = 0.007, p = 0.03), with more recent papers finding higher prevalence of psychotic disorders. There were no significant effects of age (β = 0.003, p = 0.28) or sex (β = 0.1, p = 0.41) on psychotic disorder screening rates. In overall analysis, there was significant evidence of publication bias, such that smaller samples provided higher estimates of point prevalence (see Supplemental File 3, Egger’s test for publication bias: β = 0.08 [-0.02-0.03], Z = 6.95, p < .0001).

## DISCUSSION

This is the first study to systematically review and conduct a meta-analysis on the point prevalence of psychotic experiences, CHR-P, and psychotic disorders in general psychiatric settings. Self-reported psychotic experiences were surprisingly common, identified in approximately 42% of general psychiatric settings and 45% of child and adolescent settings (90% outpatient samples). Prevalence estimates of CHR-P derived from psychiatric interviews was 26%, which was considerably higher than estimates based on a two-stage screening process (11%). Based on standardized clinical interviews for CHR-P syndromes and psychotic disorders, 26% and 6%, respectively, of treatment-seeking patients met criteria for these diagnoses, far exceeding the estimated general population rates of 3%^40^ and 1%,^102^ respectively.

The high prevalence of psychotic experiences and CHR-P in treatment-seeking samples confirms that clinical settings are risk-enriched, underscoring the need for broad and regular screening for psychotic experiences and CHR-P in these settings as a strategy for preventing both the worsening of symptoms (psychotic symptoms, comorbid conditions and related distress) and functioning and the onset of additional psychiatric conditions^103,104^ including psychotic disorders.^19^ The prevalence of CHR-P cases observed was similar to a recent meta-analysis showing a 19.2% rate in psychiatric samples.^40^ Although Salazar de Pablo and colleagues^40^ did not differentiate between types of psychiatric treatment settings, we did not find significantly different rates between general inpatient, outpatient, and child and adolescent mental health settings across all outcomes. This non-significant difference between settings could be due to the small sample size for screening inpatient and youth mental health settings; rates in these settings were likely underestimated as more acute symptoms and functional impairment are associated with higher risk of CHR-P and onset of psychosis.^102^ We did not find a difference in prevalence based on structured interview used (SIPS versus CAARMS) similar to prior meta-analysis.^40^ CHR-P rates may also be underestimated if individuals are being treated with antipsychotic medications, either for treatment of known psychotic symptoms or as adjunctive treatment for mood disorders or behavioral dysregulation. Across both psychotic experiences and CHR-P assessments, we also found a significant trend towards increased rates with more recent publications. This is in contrast with recent reports of declining rates of conversion to psychotic disorders in the CHR-P literature^19^ and may represent increasing awareness and referral to mental health services for individuals reporting psychotic experiences. In some countries, most notably Australia, public health campaigns have been conducted to increase awareness and reduce the stigma associated with psychosis.^105^ Implementation of such campaigns along with routine psychosis screening may bring more attention to these symptoms and improve early detection.

Two-stage, sequential screening could be a promising strategy for improving identification of individuals with CHR-P and psychotic disorders in general psychiatric settings, increasing rates of accurate detection compared to usual clinician referral, as demonstrated by the EDIE-NL study.^33^ About 20% of included studies in this review employed two-stage screening. With this screening strategy, 1 in 10 treatment-seeking individuals are likely to be detected for CHR-P and 1 in 25 are likely to be detected with psychotic disorders. Two-stage screening reduces likelihood of false positives, which can conserve resources by avoiding unnecessary service provision.^106,107^ Using sensitive pre-screening measures such as the Prodromal Questionnaire or PRIME Screen-Revised can also be more efficient and logistically feasible than conducting extensive CHR-P gold-standard structured clinical assessments. As an illustration, given that approximately 25 of 100 psychiatric outpatients are likely to meet CHR-P criteria and the Prodromal Questionnaire-Brief (PQ-B) has 80% sensitivity and 60% specificity^85^ in identifying likely CHR-P (based on a screening cut off of ≥ 6), using the PQ-B as a pre-screening self-report measure will reduce the number of patients needed to be assessed by half to identify individuals with true CHR-P status (positive predictive value: 40%). Thus, a 22-item questionnaire can be completed by the patient to identify potential CHR-P, in comparison with using a 1–2-hour long SIPS interview by a trained rater. Thus, based on these data, for improved identification rates and less intensive deployment of resources for screening, we recommend implementing a sequential screening process, in which a universal self-report psychotic symptom screening instrument is followed by secondary interview-based assessments in higher risk or more symptomatic populations.

A variety of public-domain, brief screening measures are good candidates for pre-screening clinically indicated psychotic symptoms (see Table 2); they have strong content and face validity, have similar psychometric properties,^85,108^ and are highly correlated with each other.^109^ Most differences in these self-report assessments are in the inclusion or exclusion of items focused on negative symptoms and other symptom clusters, the number of items, and modifications to assess a pediatric population (e.g., Prodromal Questionnaire-Brief Child Version^87^). As treatment-seeking individuals in general psychiatric settings are more likely to be experiencing more severe and impairing symptoms, it is recommended that more conservative and lower cut-off scores for identifying self-reported psychotic symptoms be used in these settings compared to cut-offs used during screening in the general or school populations.^85^ Further, given that provider and patient time burden is a significant barrier to implementing systematic screening in these settings,^110^ using very brief and easy to administer and score measures may be important for adoption. For example, the 7-item Adolescent Psychotic-Like Symptom Screener^111^ (APSS) and the 2-item psychosis screener^93^ are comprised of items with the most predictive power of other validated questionnaires (e.g., PQ-B, YPARQ-B) and point to the importance of screening for experiences of perceptual abnormalities (e.g., "Do you see things that others can’t or don’t see?") and delusions or thought disorders (e.g., "Have you ever felt that someone was playing with your mind?").^112^ More research is required to assess the feasibility and screening properties of these brief measures in general psychiatric settings specifically.

Similarly, assessing CHR-P with structured clinical interviews requires both substantial clinician time for training and establishing reliability, and a significant amount of time to complete (1-2 hours per patient interview).^18^ For a briefer CHR-P assessment, clinicians could consider using the Mini-SIPS, which takes approximately 15-20 minutes to administer and only requires a 30-minute online training to become familiar with the instrument.^97^ While the widely-used SIPS and the Comprehensive Assessment of At-Risk Mental States (CAARMS) are highly sensitive for diagnosing CHR-P and have excellent predictive validity for the conversion to psychotic disorder,^113^ there is an ongoing effort to harmonize the attenuated positive symptom ratings, severity scores, and diagnostic criteria of these two instruments into the PSYCHS.^96,114,115^ Alternatively, web-based versions of clinical interview assessments, such as the Mini-SIPS, KSADS-COMP and NetSCID, take half the time of pen-and-paper clinician assessments,^87,97^ and can be easily integrated with the increase in telehealth use in mental health services.^116^ Besides more accessible administration formats, novel screening techniques, including digital phenotyping or machine learning algorithms based on electronic health record or speech and language data may improve screening techniques.^117–119^ While this field remains nascent and such automated screening methods are not yet regularly available or integrated into clinical practice, “digital phenotyping” shows significant promise for improving screening while reducing clinician burden.

### Implications

With these meta-analytic base rates, along with the established screening properties of validated measures for detecting psychotic experiences and psychotic disorders, we can estimate the positive screen rates and resources required to assess and treat individuals with psychotic spectrum illnesses in general psychiatric settings. Distressing psychotic experiences were common, affecting two out of every five patients treated in a general psychiatric setting, without necessarily a psychotic disorder. Implementing psychosis screening should be accompanied by clinician training on how to normalize transdiagnostic psychotic experiences, and communicate about psychosis-risk and psychotic disorders,^120^ as well as feasible pathways for appropriate early intervention. Implementing routine screening with clinician training can overcome stigma and misinformation about psychotic disorders that persist even within psychiatric and mental health settings.^121^ Such training can also address knowledge gaps in terms of recognition of psychotic symptoms or disorders and both patient and provider discomfort with discussions about psychotic symptoms or psychosis-spectrum diagnoses.^110^ Systematic screening in general psychiatric settings may also help to address racial-ethnic and class disparities in diagnosis and treatment access for psychosis.^122,123^

Ideally, positive screens can be referred to specialized treatment programs for CHR-P or early psychosis; however, few coordinated specialty care programs for early psychosis are available for the number of patients with psychotic disorders. In 2018, there were 244 coordinated specialty care programs available in the United States with a capacity to serve only 8,255 patients.^124^ Thus, most patients with psychotic disorders will continue to be treated in general psychiatric settings. We anticipate that the lack of specialty care availability may reduce support for screening. However, this gap in care only further emphasizes the need to build capacity in scalable interventions and preventative approaches for CHR-P and early psychosis in general psychiatric and community care. While access to specialty treatment programs for CHR-P is limited, mental health providers in general psychiatric settings are still well-positioned to provide ongoing support and monitoring for this high-risk population.

### Limitations

This study had several limitations that generally point to an underestimation of prevalence rates of psychotic experiences and likely psychotic disorders. First, the sample was composed primarily of outpatient settings, limiting our ability to generalize these findings to inpatient samples, which may have a higher prevalence of patients with psychosis. Given the small number of inpatient samples, we did not directly compare prevalence between the two settings. Second, we did not find a significant effect of age or gender in our moderator analyses, despite known differences in age-of-onset of diagnoses.^102^ The age range in the samples was relatively restricted (78% of sample were between 14-27-years-old). The samples screened were young (mean age: 24 years, close to the mean age of onset of psychotic disorders), which may have underestimated psychotic disorders prevalence, as psychotic disorders illness typically emerge in the twenties, with a minority of patients experiencing onset of their illness even later.^102^ Third, the patients included may have been treated with antipsychotic medications which may have reduced or eliminated psychotic symptoms or under-estimated the number of individuals meeting criteria for a psychotic disorder. Fourth, heterogeneity was high across outcomes likely due to the different measures used; however, random effects analyses were conducted to account for between-sample differences, and we did not find any significant differences between types of measures used. Lastly, there was evidence of significant publication bias in the CHR-P and psychotic disorders samples. The bias in these studies was towards smaller sample sizes estimating higher prevalence rates. This publication bias suggests that smaller samples which had a lower prevalence rate may have been less likely to be published. Use of the REML method down-weights the contribution of smaller sample sizes to overall point prevalence, assuming lower reliability among smaller samples.

### Conclusions

Psychotic experiences, psychosis-risk syndromes, and psychotic disorders are common in general mental health settings and can be detected accurately and efficiently using less resource-intensive two-stage screening procedures and brief, validated screening tools. By elevating psychosis screening to the level of depression, anxiety, suicide risk, and substance use screening that is routinely implemented in general psychiatric settings, secondary prevention of adverse outcomes among individuals with psychiatric illness can be greatly enhanced. Implementing systematic psychosis screening with adequate clinician training and increased availability of evidence-based interventions for CHR-P and early psychosis in non-specialty and community settings can promote early detection and reduce the duration of untreated psychosis. Complementing psychosis screening and early intervention in the frontlines of general mental health care can prevent comorbid illnesses from developing and improve functional status and overall quality of life of the large number of patients with currently under-recognized psychosis.

## Data Availability

All data produced in the present work are contained in the manuscript.

## Acknowledgments

We are grateful to Seham Albalawi, BA, Zachary Marinov, BA, and Jeffrey Kwon, BA, for contributing to the title and abstract screening of articles for inclusion in this meta-analysis.

## Supplemental File 1

### Search Terms

**<u>Web of Science</u>**

**<u>Searched on June 29, 2023, 19:53:14 GMT</u>**

**Results: N=5525**

((TS=(psychosis)) OR (TS=(psychotic)) OR (TS=(clinical high-risk)) OR (TS=(schizophrenia))) AND ((TS=(screen*)) OR (TS=(detect*)) OR (TS=(identif*))) AND ((TS=(help-seeking population)) OR (TS=(general mental health)) OR (TS=(outpatient mental health)) OR (TS=(mental health clinic)) OR (TS=(community mental health)) OR (TS=(clinical population)) OR (TS=(referral triage)) OR (TS=(mental health services)) OR (TS=(psychiatric)) OR (TS=(counseling)) OR (TS=(counseling center))) NOT (TS=(school)) NOT (DT=(Conference Proceedings)) and 1.21 Psychiatry or 6.24 Psychiatry & Psychology or 1.100 Substance Abuse or 1.136 Autism & Development Disorders or 1.14 Nursing or 1.156 Healthcare Policy (Citation Topics Meso) and 1.21.24 Schizophrenia or 1.21.1363 Mental Health or 1.21.1828 Neuropsychiatric Disorders or 1.14.1293 Emergency Department or 1.14.724 Shared Decision Making or 1.14.763 Evidence-based Practice or 1.14.265 Nursing or 1.14.2063 Nurse Practitioner or 6.24.2075 Community Pediatrics or 1.14.363 Nursing Education or 1.156.1509 Unified Health System or 6.24.1266 Social Work Practice or 1.14.1957 Referrals or 1.14.2441 Public And National Health Services or 1.156.2181 Public Health Workforce (Citation Topics Micro)

**<u>PubMed</u>**

**<u>Searched on June 27, 2023, 17:53:27 GMT</u>**

**Results: N=553**

((("psychotic disorders"[MeSH Major Topic] OR "schizophrenia"[MeSH Major Topic] OR "psychosis"[Title/Abstract] OR "schizophrenia"[Title/Abstract]) AND ("screen*"[MeSH Major Topic] OR "early identification"[All Fields] OR "detect*"[Title/Abstract] OR "screen*"[Title/Abstract]) AND ("outpatient mental health"[All Fields] OR "mental health clinic"[All Fields] OR "community mental health"[All Fields] OR "mental health services"[All Fields] OR "help-seeking"[All Fields] OR "clinical population"[All Fields] OR "counseling"[All Fields])) NOT "school*"[All Fields]) NOT "primary care"[All Fields]

## Supplemental File 2 Screening measures for psychotic symptoms and likely psychotic disorders

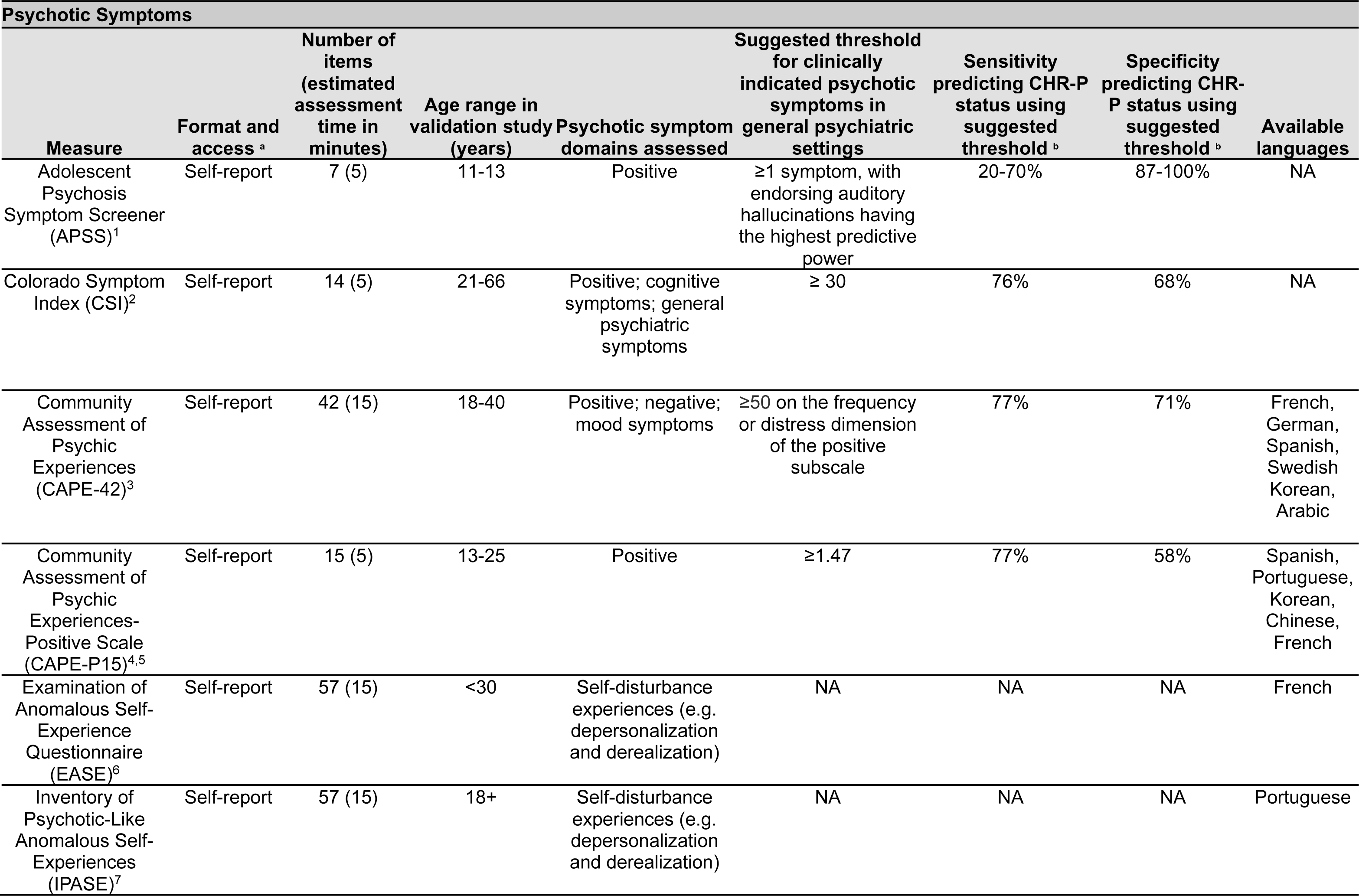
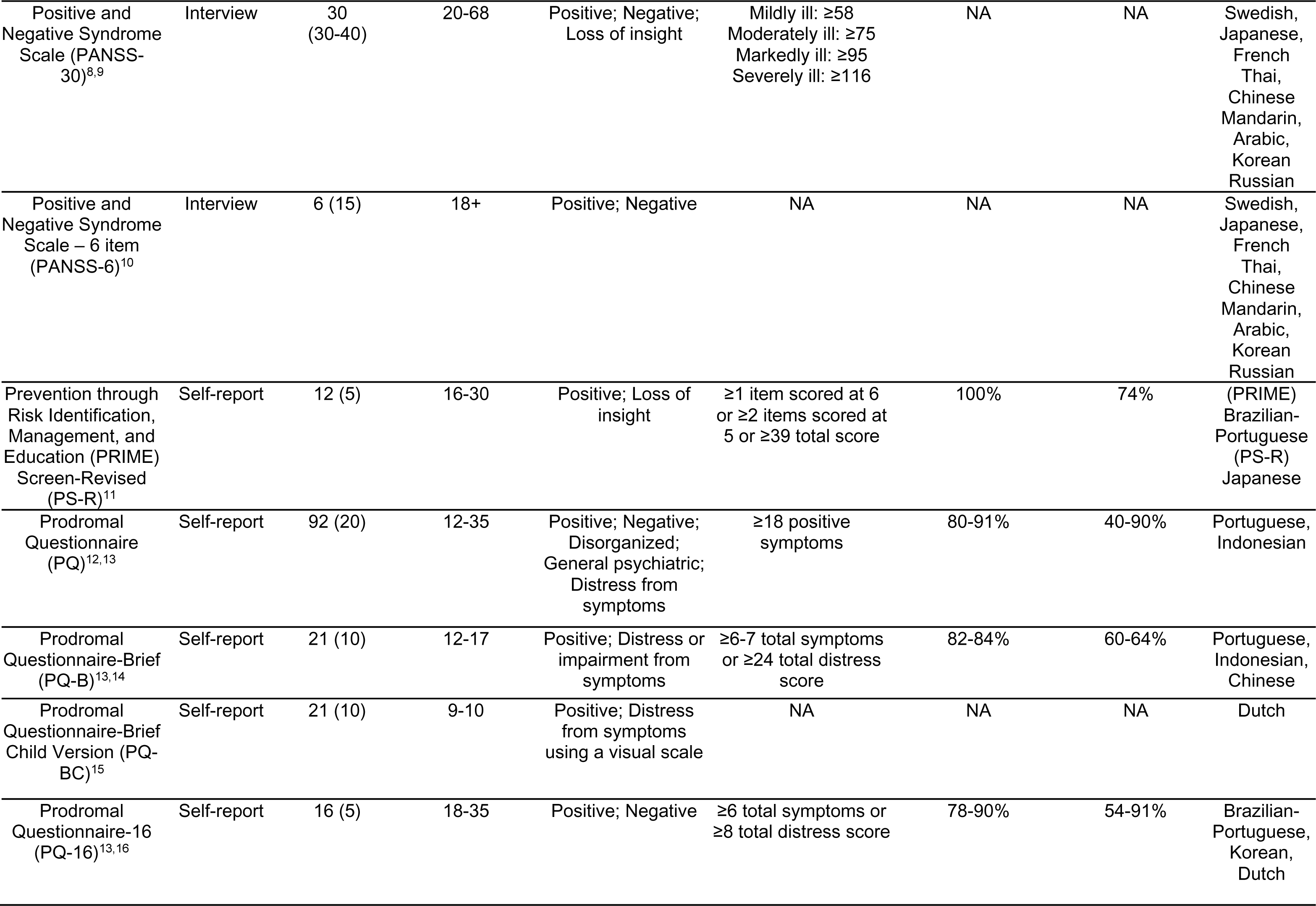
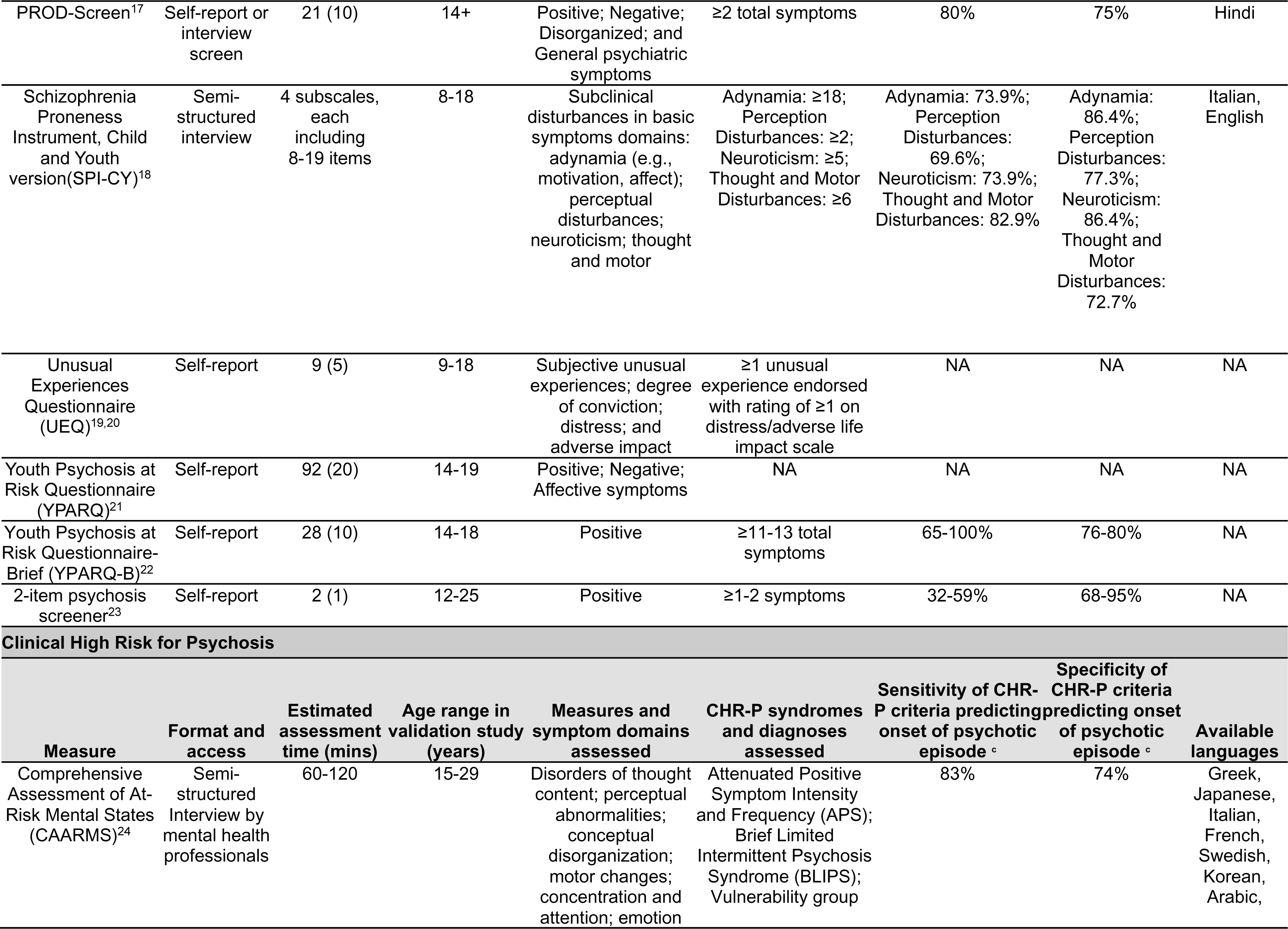
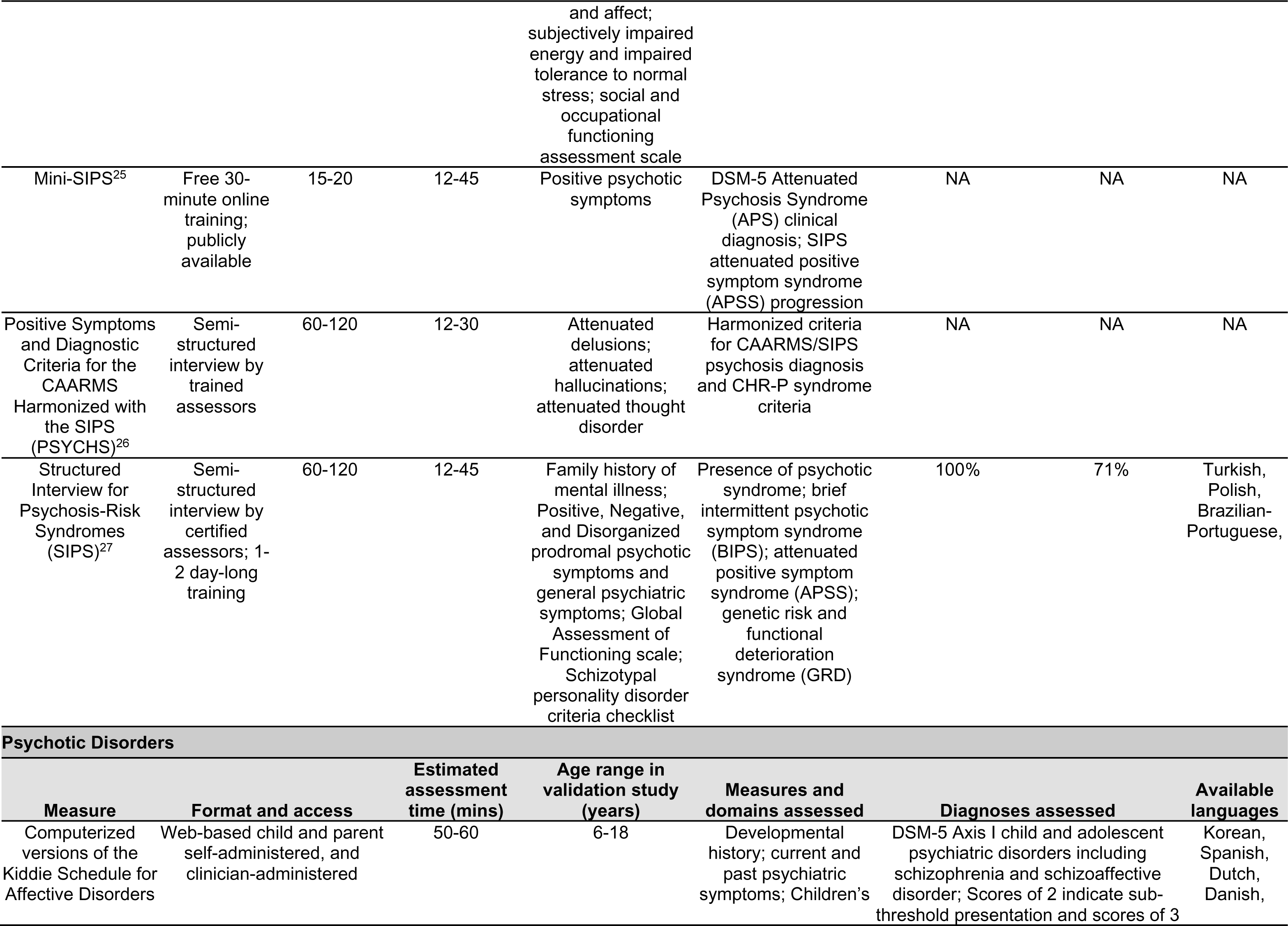
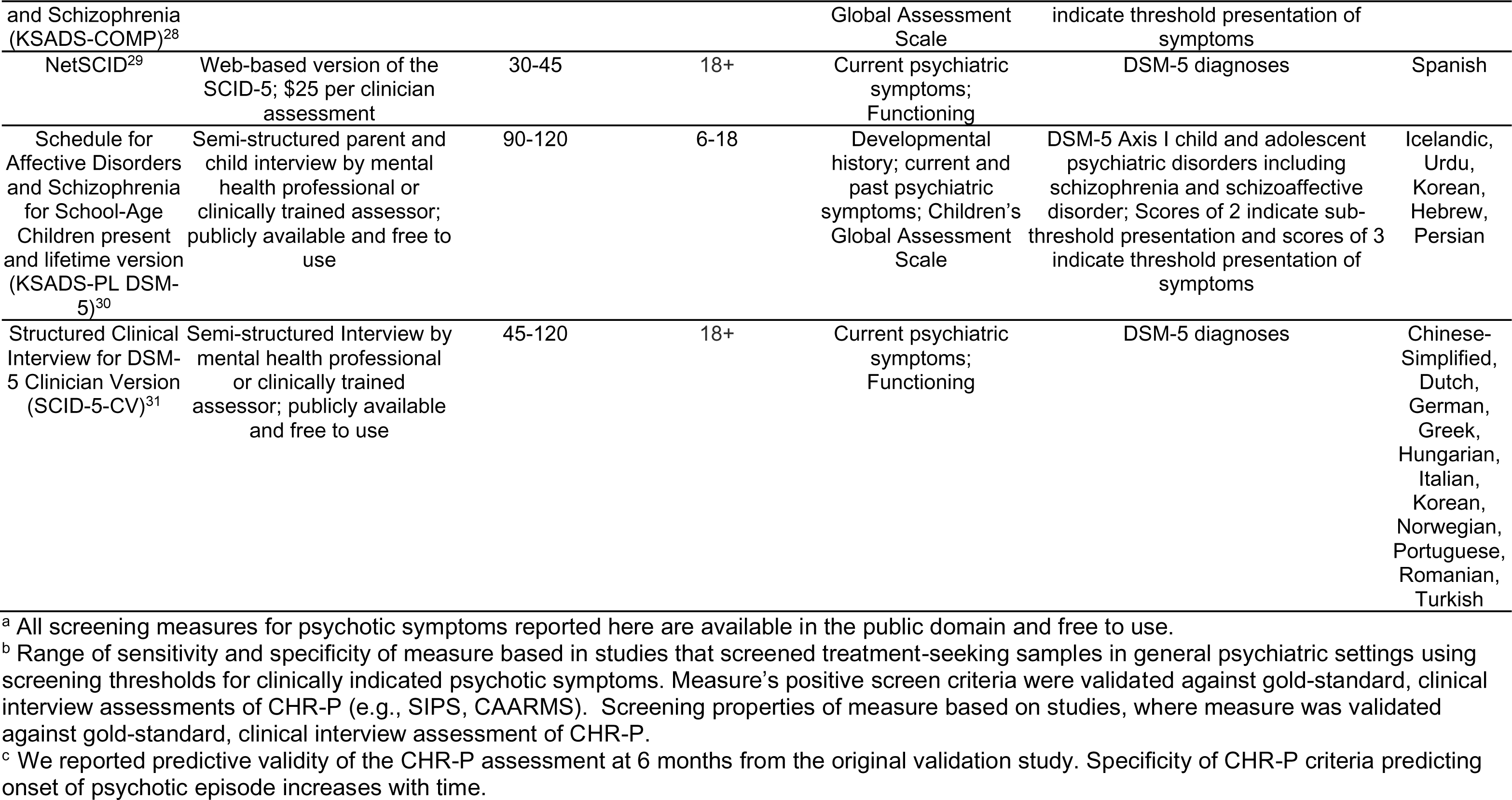

---

**Supplemental Figure 3.**
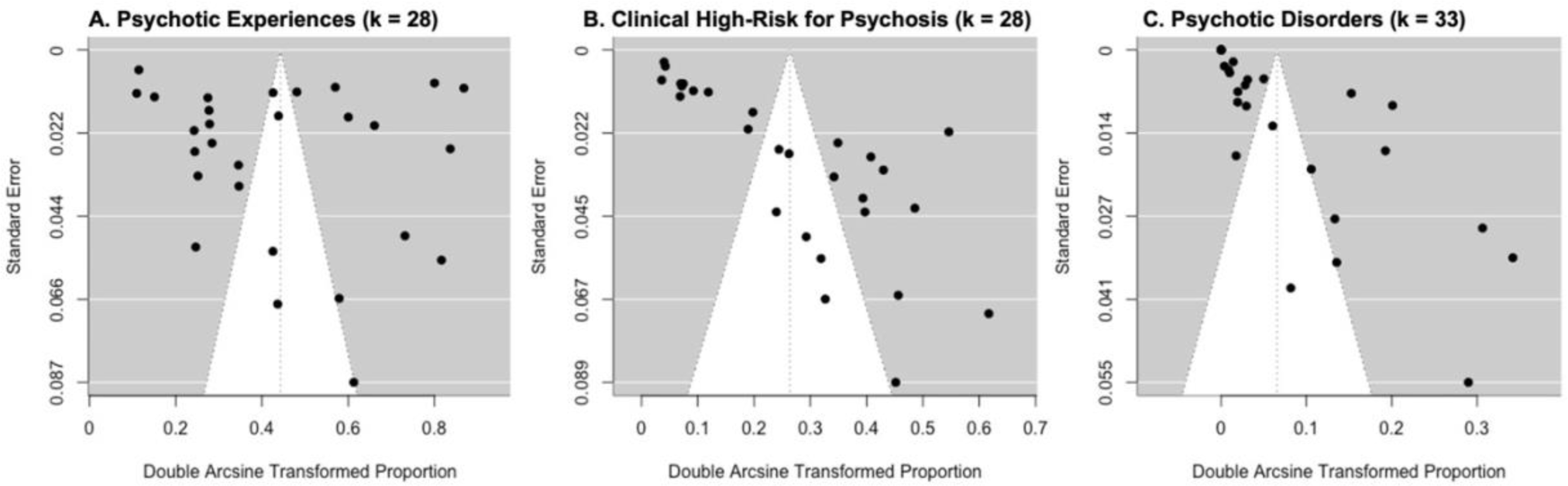
Funnel plots. Funnel plot of A) psychotic experiences; B) clinical high-risk for psychosis; and C) psychotic disorders. Each plot shows the effect size on the x-axis and the standard error on the y-axis. Vertical line is the overall estimate of the model. The white area represents a pseudo-confidence interval generated around the model estimate. Significant publication bias was identified in B) CHR-P and C) psychotic disorders with bias towards smaller samples representing larger prevalence rates.

